# Self-Management of Diabetic Patients from the Urban Areas in Bangladesh

**DOI:** 10.1101/2022.08.23.22279128

**Authors:** Md. Jubayer Hossain, Syeda Tasneem Towhid, Sumona Akter, Muhibullah Shahjahan, Tilottoma Roy, Bithi Akter, Tanjum Ahmed Nodee

**Author notes:** Corresponding author Dr. Syeda Tasneem Towhid.

## Abstract

**Background:** This study aimed to determine the knowledge about self-management and the effects of diabetes on the daily activities among adult patients with Type 1 and Type 2 diabetes in Bangladesh.

**Methodology:** A cross-sectional study was conducted from April to August 2021 on diabetic patients from Dhaka and the Rangpur district in Bangladesh. A structured questionnaire was used to collect data from study participants. This study’s researchers collected data via phone interview. The collected data was analyzed using Python (Version 3.8), Pandas, and ResearchPy (A Python package for research data analysis).

**Results:** Out of the 303 participants, all are middle-class service holders with an urban sedentary lifestyle. 69.64% and 30.3% are male and female, respectively, with 66% having post-secondary education and 35% having smoking habits. Most (68.32%) had onset of diabetic symptoms between 36-50 years, 23.43% with Type I and 41.9% with Type II diabetes. However, 34.65% of participants couldn’t answer about their ailment definitively. 22.44% of participants were on regular insulin shots within 3 months of the first diagnosis, and 90% were satisfied with managing diabetes. However, 54.46% admitted to adopting an altered lifestyle after becoming diabetic. Males and females with both types of diabetes showed significantly different comorbidities in a paired 2-tailed t-test (0.012 and 0.02 for Type I and Type II, respectively). The male participants showed higher incidences of cardiac issues, while females showed a higher propensity to hypertension. 85% of participants were confident about their medication, course of management, prognosis, diet, and lifestyle for controlling diabetes.

**Conclusions:** The disparity in the number of male and female participants and the high percentage of participants with no factual information about their prognosis need to be targeted for further study, and a patient-friendly engagement/ information dissemination action plan should be helpful.

## Introduction

Diabetes mellitus (DM), defined by a higher blood glucose level, is a metabolic complication with multiple etiologies that causes disruptions in carbohydrate, fat, and protein metabolism due to dysfunction in insulin secretion, insulin action, or both(1). This long-term high blood glucose level and the resulting metabolic deregulations are linked to secondary illness in different organ systems, including the kidneys, eyes, nerves, and blood vessels (https://idf.org/). Diabetes is a public health issue because it is prevalent in both developed and developing countries. In addition, diabetes is widely regarded as one of the leading causes of sudden illness, death, and disability worldwide(2).

According to the International Diabetes Federation (IDF), 463 million people worldwide have diabetes in 2019, nearly 20% (136 million) being 65 or older. In addition, diabetes will affect 578 million adults by 2030 and 700 million by 2045. Diabetes affected approximately 87.6 million people in South-East Asia in 2019 and is expected to increase to 152.8 million by 2045. The IDF also predicted that due to population aging, the number of people over the age of 60 who have diabetes would more than double in the next 50 years. Diabetes prevalence in Vietnam is expected to rise from 8.8 percent in 2019 to 11.3 percent in 2045, in line with the global trend(3).

A recent meta-analysis in Bangladesh, which had a population of 149.8 million people in 2011, discovered that the prevalence of diabetes among adults had increased significantly, from 4% in 1995 to 2000 and 5% from 2001 to 2005 to 9% in 2006 to 2010. According to the International Diabetes Federation, 13% of people will have diabetes by 2030(4).

A complete lack of insulin distinguishes type 1 diabetes due to the death of the pancreas’ insulin-producing beta cells. Type 2 diabetes mellitus is defined by two fundamental abnormalities. The first anomaly in developing type 2 diabetes mellitus is insulin resistance, usually compensated for by increased insulin production. Second, type 2 diabetes mellitus is caused by a malfunction in insulin secretion, which prevents it from keeping up with the increased demands imposed by the insulin-resistant condition. The proportion of children and adolescents with type 2 diabetes has increased over the last two decades. Type 1 diabetes, on the other hand, is still the most common type of diabetes(5). Diabetes mellitus is a life-long condition, and appropriate management can improve an individual’s quality of life(6). As a result, to manage diabetes, individuals and communities must first understand the disease(7). Diabetes knowledge can assist diabetic patients in avoiding the development of chronic DM commodities, which impact their quality of life.

Furthermore, people may use the information to estimate their diabetes risk, encourage them to seek appropriate treatment and care, and motivate them to manage their condition for the rest of their lives(8). Diabetes was not regarded as a severe public health concern in impoverished countries such as Bangladesh until about a decade ago, but this has rapidly changed(9). As a result, in Bangladesh, a few clinical studies on diabetes knowledge among nondiabetic and diabetic patients have been conducted(8).

This study aimed to explore diabetes-related knowledge, self-management, attitudes, and practices of type 1 and 2diabetes Mellitus populations in Bangladesh.

## Methods

A cross-sectional study was carried out from April to August 2021 of diabetic patients from Dhaka and the Rangpur district of Bangladesh towards knowledge of diabetes, self-management, and the effects of diabetes on their daily activities. The eligibility criteria for participants’ enrolment are:

1. Participants have to be Bangladeshi
2. Should be a long-term resident of Dhaka or Rangpur and be registered with the local diabetic network
3. Should be above 16 years or above
4. Should answer the complete questionnaire correctly

### Data Collection

A structured questionnaire was developed to collect information from eligible participants. The questionnaire was prepared based on a literature review of similar studies conducted in different parts of the world with some modifications in the questions based on common practice knowledge in Bangladesh. The questionnaire was separated into four sections; participants’ demographic information, self-reported diabetes type, self-management of diabetes, and the level of knowledge regarding diabetes. The data was collected through phone interviews of every participant with their consent.

## Results

### Demographic Summary

The demographic profile of the individuals in this study is shown in (Table 1). The study included 303 respondents. 211(69.6%) were male and 92 (30.36%) female. All were married and unmarried, with 286 (97.69%) and 7(2.31%). Their ages ranged from 20 to 80, with a mean age of 52.53. 157(51.82%) of the respondents were aged 51-65 years, 108(35.64%) were aged 36-50 years, 23(7.59%) were aged 66 years or over, 15(4.95%) were aged 16-35 years. The education level of this study participants, 93(30.69%) were Postgraduates, 76(25.08%) were Graduate, 34(11.22%) were undergraduates,23(7.59%) had no education, 40(13.20%) were HSC students, 37(12.21%) were SSC students. 74(24.42%) were doing public service, 41(13.53%) were retired person, 36(11.88%) were private service, 24(7.92%) were School or College teachers, and 16(5.28%) were doing police and defense service, 8(2.64%) were Bank and Financial Institution, 8(2.64%) were Doctor Physician, 7(2.31%) were private or public university teacher, 6(1.9%) were health care service provider, 5(1.65%)were Student, and 11(3.63%) were others.

**Table 1:** Demographics summary of (n = 303) respondents

| Variables | Count(n) | Percentage (%) |
| --- | --- | --- |
| **Gender** |  |  |
| Male | 211 | 69.64 |
| Female | 92 | 30.3 |
| **Age Group** |  |  |
| 16-35 | 15 | 4.95 |
| 36-50 | 108 | 35.64 |
| 51-65 | 157 | 51.82 |
| 66 and above | 23 | 7.59 |
| **Marital Status** |  |  |
| Married | 296 | 97.69 |
| Unmarried | 7 | 2.31 |
| **Education Level** |  |  |
| No Education | 23 | 7.59 |
| SSC | 37 | 12.21 |
| HSC | 40 | 13.2 |
| Undergraduate | 34 | 11.22 |
| Graduate | 76 | 25.08 |
| Post Graduate | 93 | 30.69 |
| **Occupation** |  |  |
| Public Service | 74 | 24.42 |
| Housewife | 67 | 22.11 |
| Retired | 41 | 13.53 |
| Private Service | 36 |  |
| School / College Teacher | 24 | 7.92 |
| Police and Defence Service | 16 | 5.28 |
| Bank and Financial Institution | 8 | 2.64 |
| Doctor / Physician | 8 | 2.64 |
| Govt. / Private University -<br>Teacher | 7 | 2.31 |
| Health Care Service Provider | 6 | 1.98 |
| Student | 5 | 1.65 |
| Others | 11 | 3.63 |

### Estimation of Diabetes Type

It has been observed that many people with diabetes are not sure if they have Type 1 or Type 2 diabetes. This observation prompted the creation of the questionnaire, which included several questions to help people identify their type.

(Table 2) shows the estimation of diabetes type of the respondents. The participants of this survey were asked at which age they were diagnosed with diabetes; 207 (68.32%) said 36-50 years, 60 (19.80%) said 16-35 years, 29 (9.57%) said 51-65 years,6 (1.98%) said 66 years and above, and 1(0.33%) said Under 16 years. In addition, respondents reported that 71 (23.43%) had Type 1 diabetes, 127(41.91%) had Type 2 diabetes, and 105 (34.65%) said that they didn’t know.

**Table 2:** Estimation of probable diabetes type of (n = 303) respondents

| Questions | Counts(n) | Percentage (%) |
| --- | --- | --- |
| **At which age you diagnosed diabetes?** |  |  |
| Under 16 years | 1 | 0.33 |
| 16 to 35 | 60 | 19.8 |
| 36 to 50 | 207 | 68.32 |
| 51-65 | 29 | 9.57 |
| 66 and above | 6 | 1.98 |
| **What type of diabetes do you have?** |  |  |
| Type 1 | 71 | 23.43 |
| Type 2 | 127 | 41.91 |
| Don't know | 105 | 34.65 |
| **Did you inject insulin within the first 3 months of being diagnosed?** |  |  |
| Yes (Probable Type 1) | 68 | 22.44 |
| No (Probable Type 2) | 235 | 77.56 |
| **Did you continue injecting for more than one year after you first injected insulin?** |  |  |
| Yes (Probable Type 1) | 84 | 27.72 |
| No (Probable Type 2) | 219 | 72.28 |

Using the classification of probable diabetes type based on injecting insulin within three months, 68 (22.44%) of the participants were classified as having Type 1 diabetes, and 235 (77.56%) were classified as having Type 2 diabetes. The continuation of injecting insulin for more than one year indicated that 84 (27.72%) respondents were classified as having probable Type 1 diabetes and 219 (72.28%) having possible Type 2 diabetes. Compared to the respondents who said they injected insulin within the first three months after being diagnosed with diabetes, the prevalence of Type 1 and Type 2 diabetes among them was significantly lower (Fig 1).

**Fig 1:**
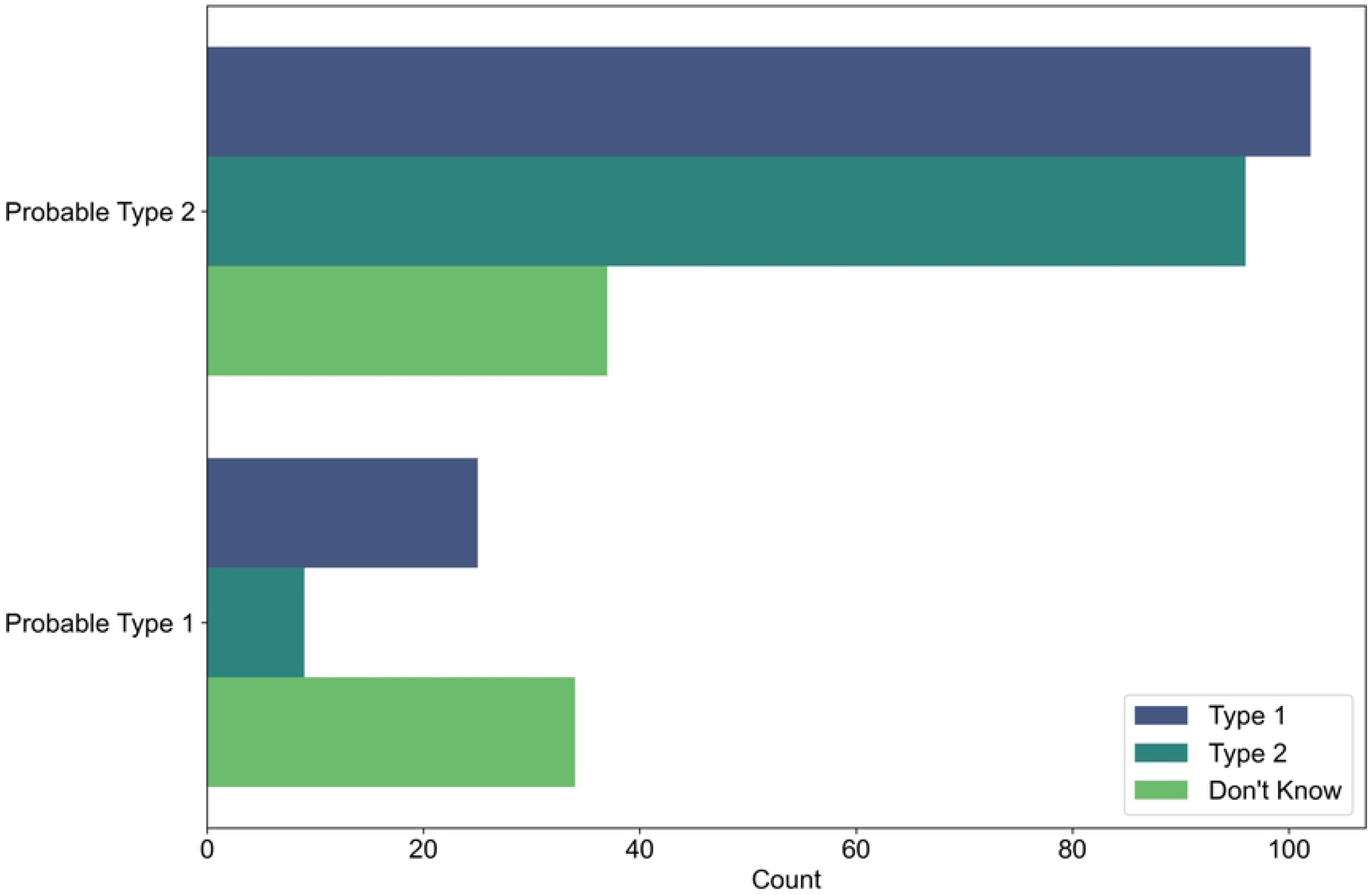
Respondent’s self-reported type of diabetes by derived diabetes (Based on injecting insulin within the first three months of being diagnosed)

Based on the continuation of insulin injection for over a year after first injected insulin, the percentage of people who could not identify their diabetes type was higher among Type 1 and 2 (Fig 2).

**Fig 2:**
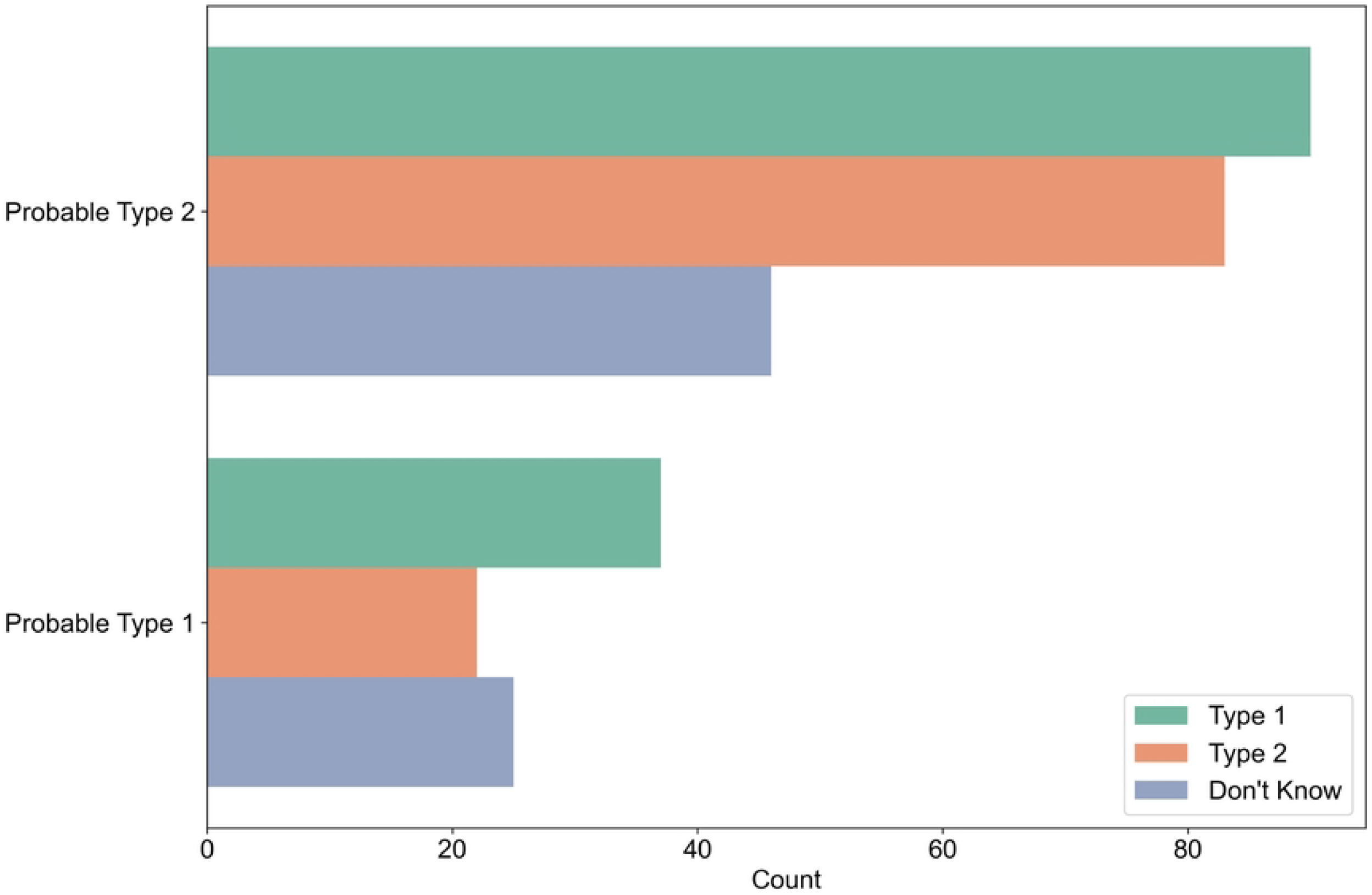
Respondents self-reported type of diabetes by derived diabetes (Based on the continuation of injecting for more than one year after first injected insulin)

Probable type 1 diabetes was diagnosed at 54.8 years; Probable type 2 was diagnosed at 51.85 years.

### Self-reported Health Status of Diabetic Patients

The self-reported health status of the respondents of this study is present in (Table 4) The study results revealed that diabetes affected the daily activities of 165 (54.46%) individuals. In addition, almost half of those with Type 2 diabetes reported that their condition affected their everyday life more than Type 1 (50.30% and 27.27%, respectively).

Participants were asked to rate their general health status in the past four weeks for the survey. They were then ranked according to how they rated their health. 134(44.22%) of the participants rated their health as ‘Good,’ 94(31.02%) as ‘Fair,’ 46(15.18%) as ‘Very Good,’ 27(8.91%) as ‘Poor,’ and 2(0.66%) as ‘Excellent.’

(Fig 3) shows that people with Type 1 said their health was good, while those with Type 2 stated it was fair. Participants aged 16 to 35 said they had an excellent or acceptable rating. However, they also said that they had a good or poor rating. The participants with the highest rating were those 36 to 50, and the lowest were those under 16 years old (Fig 3).

**Fig 3:**
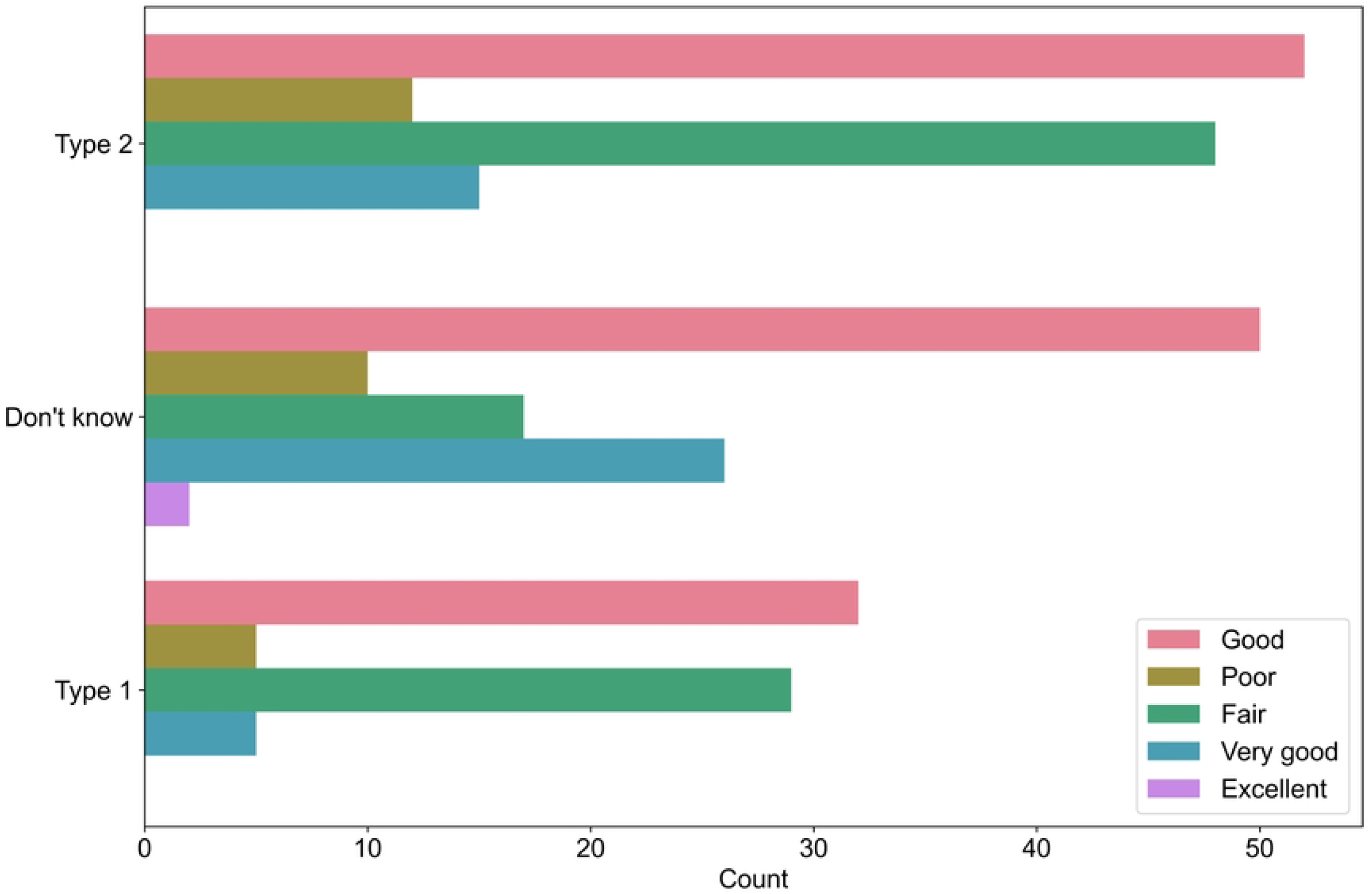
Respondent’s self-reported health status by diabetes type.

(Table 3) shows that 187(61.72%) have a family history of diabetes, and 134(44.22%) stayed in hospital overnight. The reasons for the most recent hospital admission were diabetes (11, 3.63%), Both diabetes and symptoms related to Diabetes (86, 28.38%), symptoms unrelated to diabetes (3, 0.99%), and Something else(133, 43.89%). On the other hand, 70(23.10%) did not recently stay in hospital. 136(44.88%) of the participants go to a hospital or clinic to check up on diabetes, 67(22.11%) doctor’s chamber, 54(17.82%) at home, 44(17.82%) local pharmacy, and 2(0.66%) others. Thus, most of the 244(80.53%) check-up diabetes three or more times in the last 12 months, 37(12.21%) twice, 8(2.64%), and 1(0.33%) did not check-up diabetes.

**Table 3:** Self-reported health status of (n = 303) diabetic patients

| Questions | Count | Percentage |
| --- | --- | --- |
| **Overall health in the past 4 weeks?** |  |  |
| Good | 134 | 44.22 |
| Fair | 94 | 31.02 |
| Very Good | 46 | 15.18 |
| Poor | 27 | 8.91 |
| Excellent | 2 | 0.66 |
| **Does diabetes affect day to day activities?** |  |  |
| Yes | 165 | 54.46 |
| No | 138 | 45.54 |
| **Family history of diabetes** |  |  |
| Yes | 187 | 61.72 |
| No | 116 | 38.28 |
| **Whether stayed in hospital overnight** |  |  |
| Yes | 134 | 44.22 |
| No | 169 | 55.78 |
| **Reason for most recent stay in hospital** |  |  |
| Diabetes | 11 | 3.63 |
| Both diabetes and symptoms -<br>related to diabetes | 86 | 28.38 |
| symptom unrelated to<br>diabetes | 3 | 0.99 |
| Something else | 133 | 43.89 |
| not stayed | 70 | 23.1 |

| **Where do you go for diabetes check-up** |  |  |
| --- | --- | --- |
| Hospital/clinic | 136 | 44.88 |
| Doctor's chamber | 67 | 22.11 |
| At home | 54 | 17.82 |
| Local pharmacy | 44 | 17.82 |
| Others | 2 | 0.66 |
| **Number of diabetes check-ups in the last 12 month** |  |  |
| Three or more time | 244 | 80.53 |
| Twice | 37 | 12.21 |
| Once | 8 | 2.64 |
| None | 1 | 0.33 |

**Table 4:** Self-management of diabetes of (n=303) participants.

| Questions | Count | Percentage |
| --- | --- | --- |
| How do you control your diabetes now? |  |  |
| Insulin | 83 | 26.6 |
| Tablets | 186 | 59.6 |
| Diet | 163 | 52.2 |
| Physical Activity | 136 | 43.6 |
| Others | 42 | 13.5 |
| **Do you take any medication for any other condition?** |  |  |
| Yes | 230 | 75.91 |
| No | 67 | 22.11 |
| Don't Know | 6 | 1.98 |
| **What type of medication do you take?** |  |  |
| Tablets for high blood pressure | 109 | 34.9 |
| Tablets for high cholesterol | 66 | 21.2 |
| Tablets for heart disease | 99 | 31.7 |
| No medication | 73 | 23.4 |
| Others | 106 | 34 |
| **How often do you test your own blood glucose level?** |  |  |
| Less than once | 236 | 77.89 |
| Once a Day | 22 | 7.26 |
| 4 or more times a day | 3 | 0.99 |
| 2 or 3 times a day | 2 | 0.66 |
| Never | 40 | 13.2 |

### Knowledge and attitudes regarding diabetes among diabetic patients

The results of knowledge and attitudes toward diabetes are presented in (Table 5). Most respondents knew when to take medication, but 11.12% did not know. The respondents also showed a positive attitude toward medicine. In addition, most of them were aware of the eating habits to control diabetes (Table 5). Among the participants, 65.02% were non-smokers, and 34.98% were smokers.

**Table 5:** Knowledge and attitudes regarding diabetes among diabetic patients

| **Questions** | **Count** | **Percentage** |
| --- | --- | --- |
| **Do you know enough about when to take your medication?** |  |  |
| Yes | 258 | 85.15 |
| No | 11 | 11.22 |
| I would like to know | 34 | 3.63 |
| **Do you know enough about what you should eat to help you manage your diabetes?** |  |  |
| Yes | 267 | 88.12 |
| No | 10 | 3.30 |
| I would like to know | 26 | 8.58 |
| **Do you know about the role of physical activity in managing your diabetes?** |  |  |
| Yes | 270 | 89.11 |
| No | 13 | 4.29 |
| I would like to know | 20 | 6.60 |
| **Do you smoke?** |  |  |
| Yes | 106 | 34.98 |
| No | 197 | 65.02 |

## Discussion

Over 8.3 million diabetic citizens in Bangladesh are plagued with health problems such as diabetic foot infection, diabetic retinopathy, chronic kidney disease, and stroke(10), preventable consequences of controlled lifestyle and medication. Awareness and a proactive lifestyle are pre-requisite in keeping diabetes under control. This study targets to assess the level of awareness among patients with active diabetes in Bangladesh, so an effective awareness strategy can be formulated to encounter any knowledge gap among the patients and their immediate family members. In this study, 69.64 % of participants were male, and 30.36% were female. The literacy level was primarily post-secondary (66%) and below-secondary (34%). All participants confessed to a sedentary urban lifestyle, 35% were smokers, and 68.32% reported onset of diabetic symptoms between 36-50 years of age. 23.43% were diagnosed with type I diabetes, 41.9% have type 2 diabetes, and 34.65% are unaware of the type of diabetes they have. 22.44% of participants were on insulin within the first 3 months of diagnosis, and 54.46% adopted an altered lifestyle to assume daily jogging and a low-sugar diet. 91% of the patients were satisfied with the prognosis and service provided, while 9.8% expressed dissatisfaction over how diabetes was managed. The rate of severe outcomes from diabetic syndrome was alarming: 76% of the participants reported chronic conditions like hypertension, high cholesterol, and cardiac ailment. There was no significant statistical difference between the severity of symptoms between genders regarding type I and II diabetes or age-of-onset symptoms (Fig 4).

**Fig 4:**
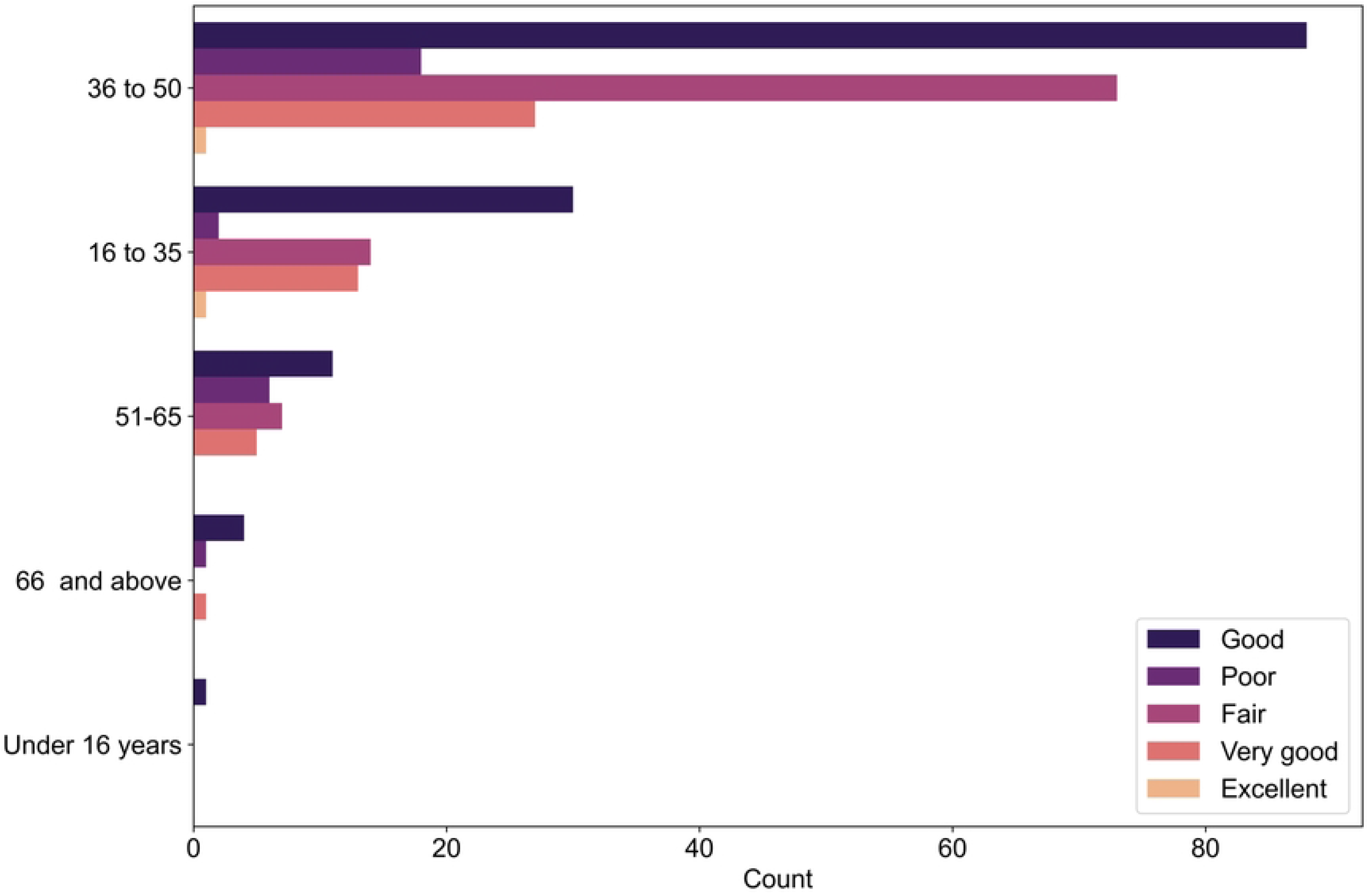
Respondent’s self-reported health status by age.

However, males and females suffer from significantly different comorbidities (p=0.12 and 0.02 for paired 2-tailed T-test regarding type I and type II diabetes, respectively). Male participants had higher incidences of cardiac issues, while females had higher hypertension. 85% of participants were confident about their prognosis, medication, diet, and lifestyle, while 15% were fully-dependent on their primary caregivers for diabetes management. Our study lacks some helpful information provided by other groups working on the same problem. Islam et.al. (2021) reported many pre-diabetic individuals in the study population, saying only one-third of participants received treatment regularly. That rate of other predisposing factors to morbidities such as obesity and hypertension was high among the affluent quarter of the society. Mohiuddin (2019) elucidated the disparity among urban and rural diabetic patients regarding healthcare access. Rahman et al. (2015) presented a similar study where age-standardized prevalence and difference were evident in diabetes management; the low-income people in Northwest Bangladesh were the most deprived of diabetes medication. Talukder and Hossain (2020) generated a two-tier logistic regression model to show females were more likely to develop diabetes mellitus, and 62% of affluent quarter get this form of diabetes. The average cost of diabetes management per head was USD 864.7 monthly.

In contrast, the mean monthly income from the same year was USD 1131(11). Hospitalization from diabetes was 4.2 times higher than non-diabetic people. Female patients spend more money on diabetes management per month. WHO Annual report (2016) stressed the necessity to formulate national guidelines such as a federal response to diabetes, evidence-based national guidelines, an operational action plan to reduce obesity, standard criteria for referral to high-level medical care, and a national risk factor survey. Taken together, our project and other available information resonate with the need for more effort on national and population levels to control and manage diabetes.

## Data Availability

The dataset used and analysed during the study is available from the corresponding author and ethical review committee.

## Ethics

This study adhered to the most significant ethical standards imaginable, and participants gave informed consent. Informed consent was also obtained from the guardian of each participant under 18 years of age. The Helsinki Declaration was observed in all procedures. Anonymity and confidentiality were maintained. Anonymity and confidentiality were maintained. We obtained ethical approval for this study from the Ethical Review Committee of CHIRAL Bangladesh (Reference No: CHIBAN21MAR2021-0003)

## Consent for publication

The authors declare that they have no known competing financial interests or personal relationships that could appear to influence the publication of this research output.

## Availability of data and materials

The dataset used and analyzed during the study is available from the corresponding author

## Authors’ contributions

Conceptualization: Md. Jubayer Hossain

Data curation: Md. Jubayer Hossain, Sumona Akter, Muhibullah Shahjahan, Tilottoma

Roy, Bithi Akter, Tanjum Ahmed Nodee

Formal analysis: Md. Jubayer Hossain

Investigation: Md. Jubayer Hossain, Dr. Syeda Tasneem Towhid

Methodology: Md. Jubayer Hossain

Project Administration: Md. Jubayer Hossain, Dr. Syeda Tasneem Towhid

Resources: Md. Jubayer Hossain

Software: Md. Jubayer Hossain

Supervision: Dr. Syeda Tasneem Towhid

Validation: Md. Jubayer Hossain, Dr. Syeda Tasneem Towhid

Writing – Original Draft Preparation: Md. Jubayer Hossain, Syeda Tasneem Towhid, Sumona Akter, Muhibullah Shahjahan

Writing – Review & Editing: Md. Jubayer Hossain, Dr. Syeda Tasneem Towhid, Sumona Akter, Muhibullah Shahjahan, Tilottoma Roy, Bithi Akter, Tanjum Ahmed Nodee

All authors have read and approved the manuscript.

